# Within-host dynamics of antiviral treatment for SARS-CoV-2 infection

**DOI:** 10.1101/2024.05.31.24308284

**Authors:** Lea Schuh, Peter V. Markov, Ioanna Voulgaridi, Zacharoula Bogogiannidou, Varvara A. Mouchtouri, Christos Hadjichristodoulou, Nikolaos I. Stilianakis

**Author notes:** corresponding authors (L.S.), (N.I.S.).

## Abstract

The effectiveness of antiviral treatment with remdesivir against COVID-19 has been investigated in clinical trials suggesting earlier recovery. However, this effect seems to be rather modest. In this study, we assessed the clinical course of SARS-CoV-2 infections in 369 COVID-19 individuals across a spectrum of illness severities, including both untreated individuals and individuals who received antiviral treatment with remdesivir. Moreover, using a process-based mathematical model, we quantified and analyzed the within-host infection dynamics of 69 untreated and 19 antiviral-treated individuals. For untreated individuals, we found that those hospitalized exhibit significantly lower levels of early immune response and higher cumulative viral loads than those who were not. For treated individuals, we found that those who died were on average hospitalized later after symptom onset than those who survived, underscoring the importance of early medical intervention for severe COVID-19. Our model estimates a rather limited antiviral activity of remdesivir and, consequently, comparable viral load dynamics between individuals responding and not responding to antiviral treatment. Our results provide valuable insights into the clinical course of COVID-19 during antiviral treatment with remdesivir and suggest the need for alternative treatment regimens.

## INTRODUCTION

Acting as an adenosine triphosphate analogue, remdesivir is meant to impede RNA-dependent RNA polymerase activity, a crucial step in viral replication. The interference with this biological mechanism is expected to significantly affect the viral dynamics of severe acute respiratory syndrome coronavirus type 2 (SARS-CoV-2) and is the basis for the desired clinical effects of remdesivir. The effectiveness of remdesivir as an antiviral in individuals infected with SARS-CoV-2 has been investigated in clinical trials, with conflicting reports regarding its impact on important clinical outcomes such as patient mortality, duration of hospital stay, or time to recovery [1–7]. Despite equivocal evidence, remdesivir has been authorized for emergency use throughout the European Union and other countries including the United States, Australia, and Japan. Understanding within-host viral load dynamics during the course of infection under antiviral treatment can provide insights into the mechanisms of drug action and the relationships between the process of infection and the above-mentioned clinical outcomes [8]. So far, studies evaluating the effectiveness of remdesivir have predominantly focused on the clinical symptoms of individuals [1,3,5] or used sparse viral load data [9–11]. By coupling mechanistic modeling and viral load measurements during the course of infection, viral dynamics can be quantified at the level of the individual [12–15], providing powerful means to evaluate the individual-specific effectiveness of COVID-19 treatments on infection dynamics [16].

Here, we present a cohort of 369 individuals with COVID-19, spanning a broad range of disease severities, including both individuals treated with the antiviral drug remdesivir and untreated individuals. Leveraging individual viral load data and process-based mathematical modeling, our study investigates poorer immune response and prolonged infections among the untreated hospitalized individuals. Among treated individuals, we examined the effects of hospitalization and antiviral treatment with remdesivir on survival. Furthermore, we estimated the effectiveness of antiviral treatment and compared viral load dynamics between individuals responding and not responding to treatment. Our analysis provides a comprehensive multi-parameter profile of SARS-CoV-2 infections across different disease severities, examining the antiviral effects of remdesivir on viral load dynamics.

## RESULTS

### Description of the study cohort

Between October 2020 and September 2022, we recruited 369 individuals at the emergency department of the public hospital in Larissa, Greece, reporting symptoms consistent with COVID-19 and/or a history of close contact with a confirmed COVID-19 case and whose naso-pharyngeal samples tested positive for SARS-CoV-2 (Figure 1).

**Figure 1.**
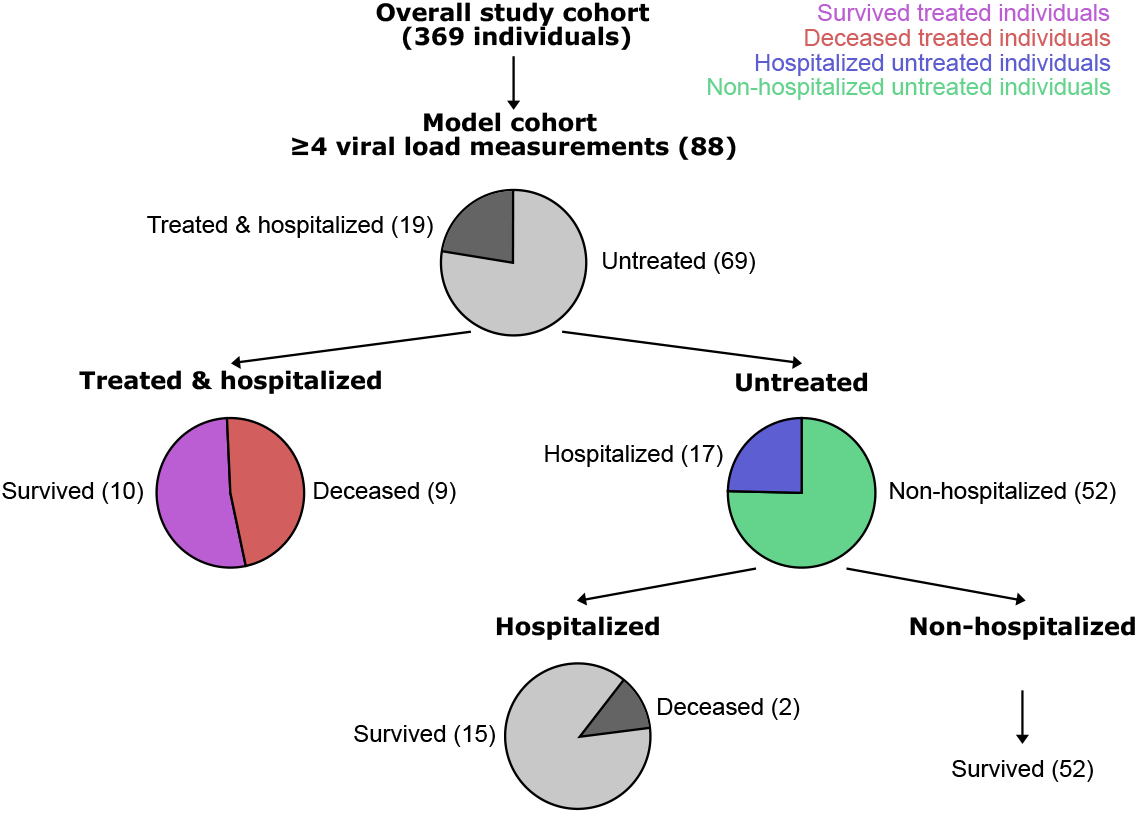
Schematic presentation of the study cohort and its subsets. Overall, 369 individuals participated in the study. Viral load measurements were taken on at least 4 different days for 88 individuals (model cohort). Of these 19 individuals were treated and hospitalized. Of these in turn 10 survived (purple) and the other 9 died from COVID-19 (red). Of the 69 untreated individuals, 17 were hospitalized (blue) and 52 were non-hospitalized (green). The proportions of the different subsets are visualized by pie charts.

Of these 205 (56%) were women and 164 (44%) were men (see full cohort description in Extended Data Figure 1 and Supplementary information). The median age of the cohort was 49 years (range 3-99 years). Most infections were primary (96%) and less than half of the cohort was previously vaccinated (44%). For our analysis we focused on a subset of 88 individuals for whom viral loads were measured on at least 4 different days through reverse transcriptase polymerase chain reaction (RT-PCR) targeting three SARS-CoV-2 specific genetic regions: ORF1ab, N, and S. We refer to this subset of individuals as the “model cohort”, which includes 19 individuals who were hospitalized and treated with antiviral remdesivir, referred to as “treated individuals”, and 69 individuals who were not treated with antiviral remdesivir, referred to as “untreated individuals”.

### Model fits to viral load dynamics in untreated individuals

To quantify the infection dynamics in untreated individuals, we fitted a within-host process-based mathematical model to viral loads derived from the observed and measured cycle threshold values for all untreated individuals (Figure 2, Extended Data Figures 2 and 3, and Extended Data Table 1).

**Figure 2.**
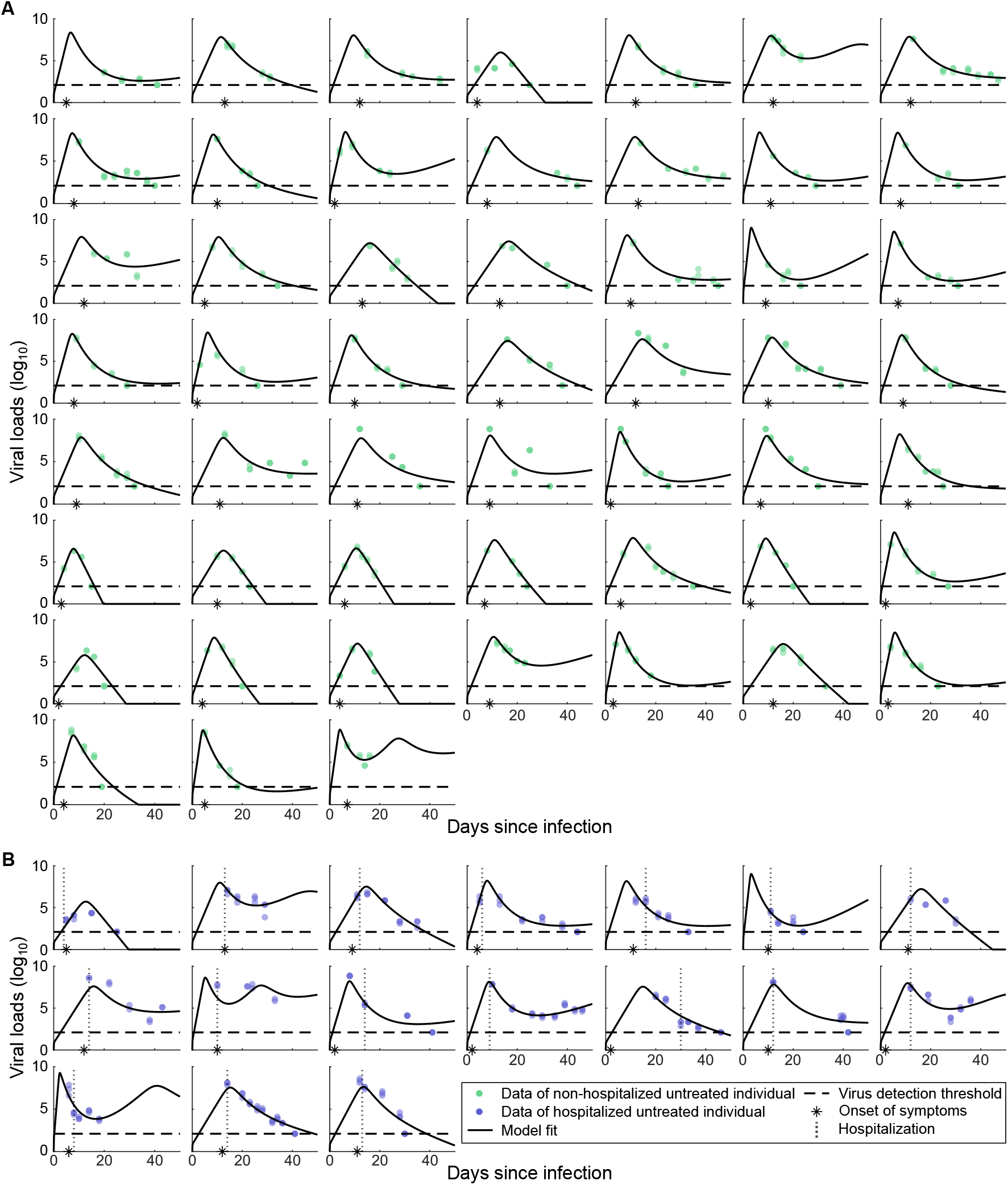
Model fits and individual level viral load measurements of untreated individuals. (A) Model fits (black line) to viral loads (VL, log_10_) of 52 non-hospitalized untreated individuals (green dots). The asterisk denotes the reported day of symptom onset. The horizontal dashed line represents the detection threshold (DT) of the RT-qPCR method to detect viral load within the sample (DT = 2.1 log_10_ VL). Dots on the dashed line denote negative test results, likely measurements below the DT. (B) Model fits (black line) to viral loads (log_10_) of 17 hospitalized untreated individuals (blue dots). The day of hospitalization is highlighted by a vertical dotted line. Symbols same as in (A).

The within-host model describes the dynamics of susceptible and infected nasal epithelial cells, free virus, and the immune response during the course of a SARS-CoV-2 infection [15]. To capture heterogeneities between individuals, we estimated three model parameters at the individual level: the incubation period (the period from infection to onset of symptoms), the viral production rate constant, and the activation rate constant of the immune response (Extended Data Table 1). Overall, the model captures the observed viral load dynamics for both non-hospitalized and hospitalized untreated individuals well (Figure 2), quantifying the nuanced dynamics of mild to severe SARS-CoV-2 infections.

### Weaker early immune response and prolonged infections in hospitalized individuals

In order to compare severe and mild COVID-19 in untreated patients, we looked at an array of dynamic features in hospitalized (severe) and non-hospitalized (mild) untreated individuals. Specifically, we compared the estimated and predicted values of timing features of the infection (Figure 3A and 3B), viral loads (Figure 3C and 3D), and immune response features (Figure 3E and 3F), as well as the estimated model parameters between these two groups (Figure 3G) (Extended Data Tables 2 and 3).

**Figure 3.**
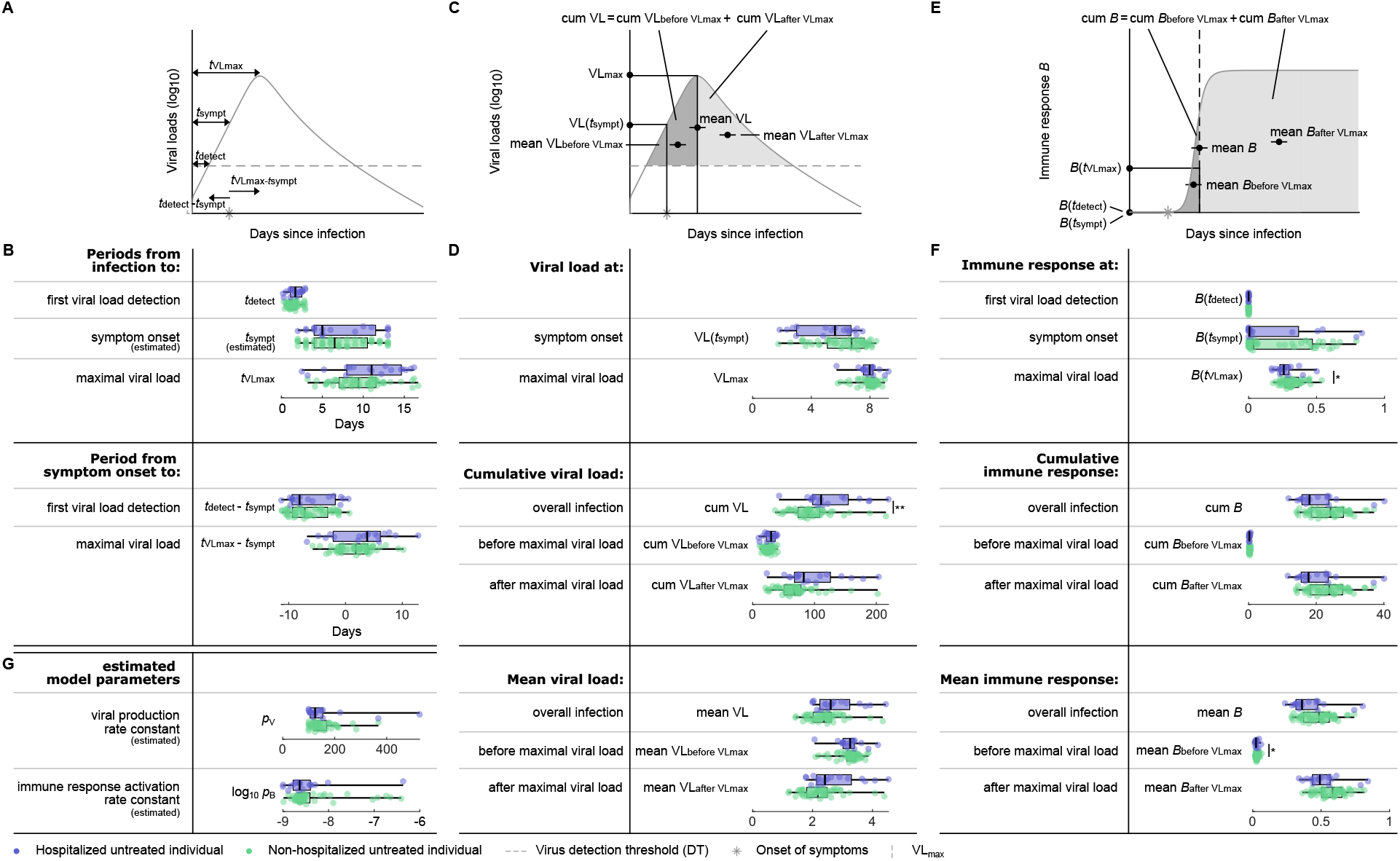
Clinical/virological timing, viral load, immune response features and model parameters for hospitalized and non-hospitalized untreated individuals. (A) Schematic presentation of timing features. (B) The estimates and predictions of the timing features in A for hospitalized (blue) and non-hospitalized (green) untreated individuals. The boxplots represent the median, lower and upper quartiles, and minimum and maximum values. All features are predictions, if not specified differently. (C) Schematic presentation of viral load features. (D) The individual predictions of the viral load features from C. (E) Schematic presentation of immune response features. (F) The individual predictions for the immune response features in E. (G) Estimated rates constants.

The majority of the investigated features and estimated model parameters quantifying the viral load dynamics did not exhibit statistically significant differences between mild and severe COVID-19 cases. However, the predicted cumulative detectable viral load over the course of an infection, represented by the area enclosed between the predicted viral load function and the determined detection threshold, is significantly greater in hospitalized untreated individuals (Figure 3D). Increased predicted cumulative detectable viral loads could be a result of either increased viral loads or prolonged infections. As not all individuals tested negative for SARS-CoV-2 after 50 days of predicted infection, a direct quantification and comparison of the infection duration is challenging. Instead, we quantified and compared the predicted mean viral loads during the infection course for hospitalized and non-hospitalized untreated individuals and did not find statistically significant differences between them. Overall, this suggests that hospitalized untreated individuals with severe COVID-19 likely demonstrate prolonged infections in comparison to non-hospitalized untreated individuals with mild COVID-19. Moreover, our analysis revealed a significantly decreased predicted immune response at maximal viral load and a significantly decreased predicted mean immune response during early infection for hospitalized untreated individuals (Figures 3F and Extended Data Figure 3), indicating a weaker immune response activation in this group. Interestingly, the weaker early immune response and prolonged infections in hospitalized untreated individuals with severe COVID-19 do not appear to depend solely on either the viral production rate constant or activation rate constant of the immune response (Figure 3G).

### Model fits to viral infection dynamics of treated individuals

We extended the within-host model [15] to investigate the viral infection dynamics of individuals receiving antiviral treatment with remdesivir. To incorporate treatment effects into the within-host model, we allowed for a reduced viral production rate throughout the duration of treatment (Extended Data Figure 2). Recognizing the variability in remdesivir’s effectiveness among individuals, we estimated it at an individual level and fitted the within-host model accounting for treatment to the viral load data of all treated individuals (Figure 4, Extended Data Figures 2 and 4, and Extended Data Table 1).

**Figure 4.**
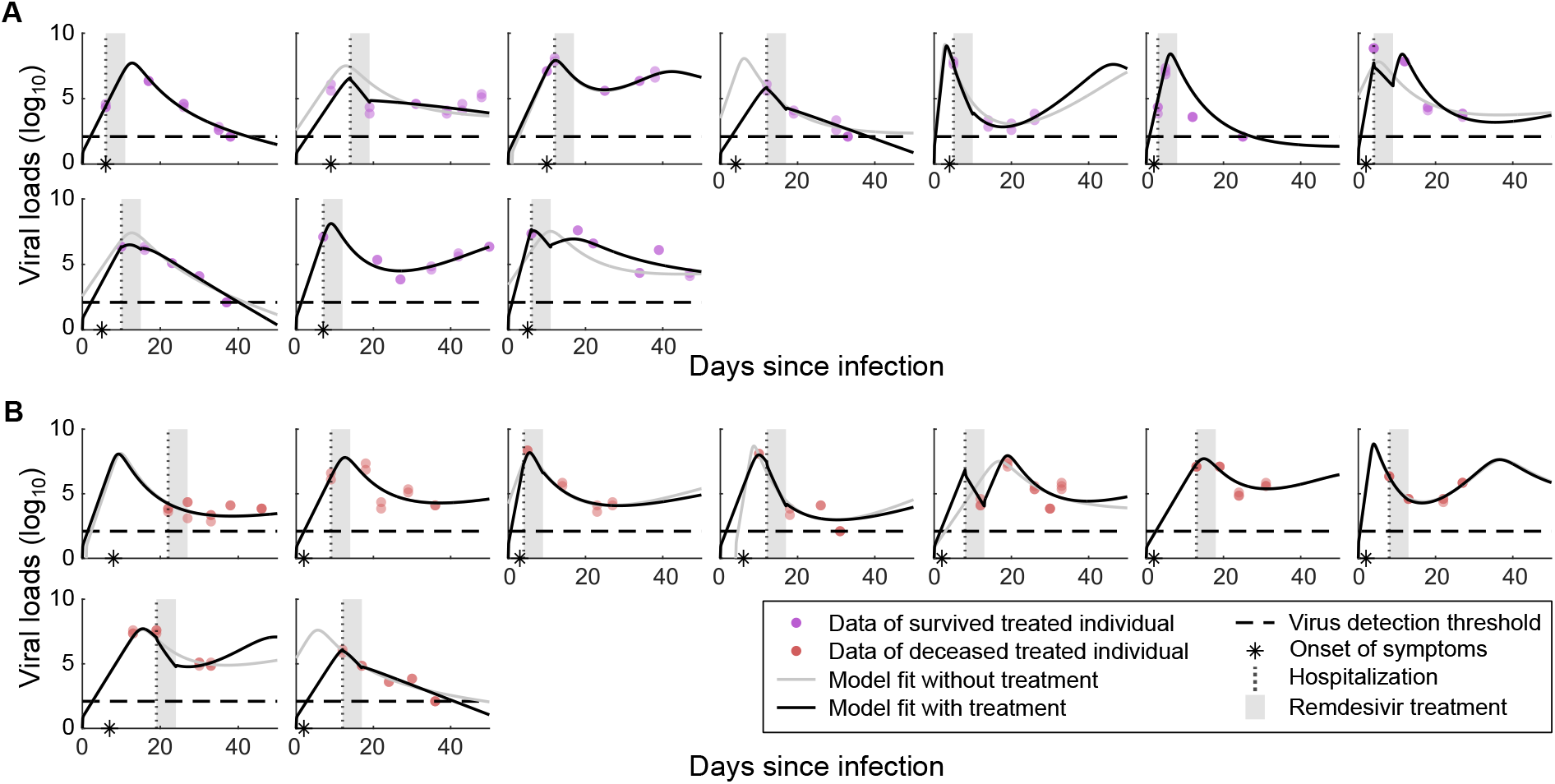
Model fits and individual level viral load dynamics of remdesivir-treated individuals. (A) Model fits with treatment (black line) and without treatment (gray line) to viral loads (log_10_) of 10 treated individuals that survived the infection (purple dots). A vertical dotted line highlights the day of hospitalization and the asterisk denotes the reported day of symptom onset. The gray area highlights the five-day period of remdesivir treatment. During that period, the viral production rate p_V_ is decreased by a factor (1-α), where α is the proportion of reduced viral production due to treatment, termed treatment effectiveness. The horizontal dashed line represents the detection threshold (DT) of the RT-qPCR method to identify virus in the sample (DT = 2.1 log_10_ VL). Dots on the dashed line represent negative test results, likely measurements below the DT. For some individuals the model fits with treatment and without treatment are overlapping. (B) Model fits with treatment (black line) and without treatment (gray line) to viral loads (log_10_) of 9 treated individuals that died from COVID-19 (red dots). Symbols same as in (A).

The model incorporating treatment effects described the observed viral loads for treated individuals well, capturing and quantifying the dynamics of both survived and deceased treated individuals. To assess the added explanatory value of incorporating treatment effects into the model, we also fitted the model without treatment to the viral load data of the treated individuals. Quantitative evaluation using the Bayesian Information Criterion indicated that, in general, the model without treatment is sufficient to describe the viral load data, with only one of the 19 individuals exhibiting a considerably improved fit with the inclusion of treatment in the model (Extended Data Table 4). This model comparison suggests that in most cases antiviral treatment with remdesivir may have a limited impact on viral load dynamics.

### Early hospitalization after symptom onset is crucial for patient survival

Focusing on the treated individuals, we compared the previously defined dynamical features, i.e., timing (Figure 5A and 5B), viral load (Figure 5C and 5D), and immune response (Figure 5E and 5F) features, and the estimated model parameters (Figure 5G) (Extended Data Tables 3 and 5) between the treated individuals that survived and those that died.

**Figure 5.**
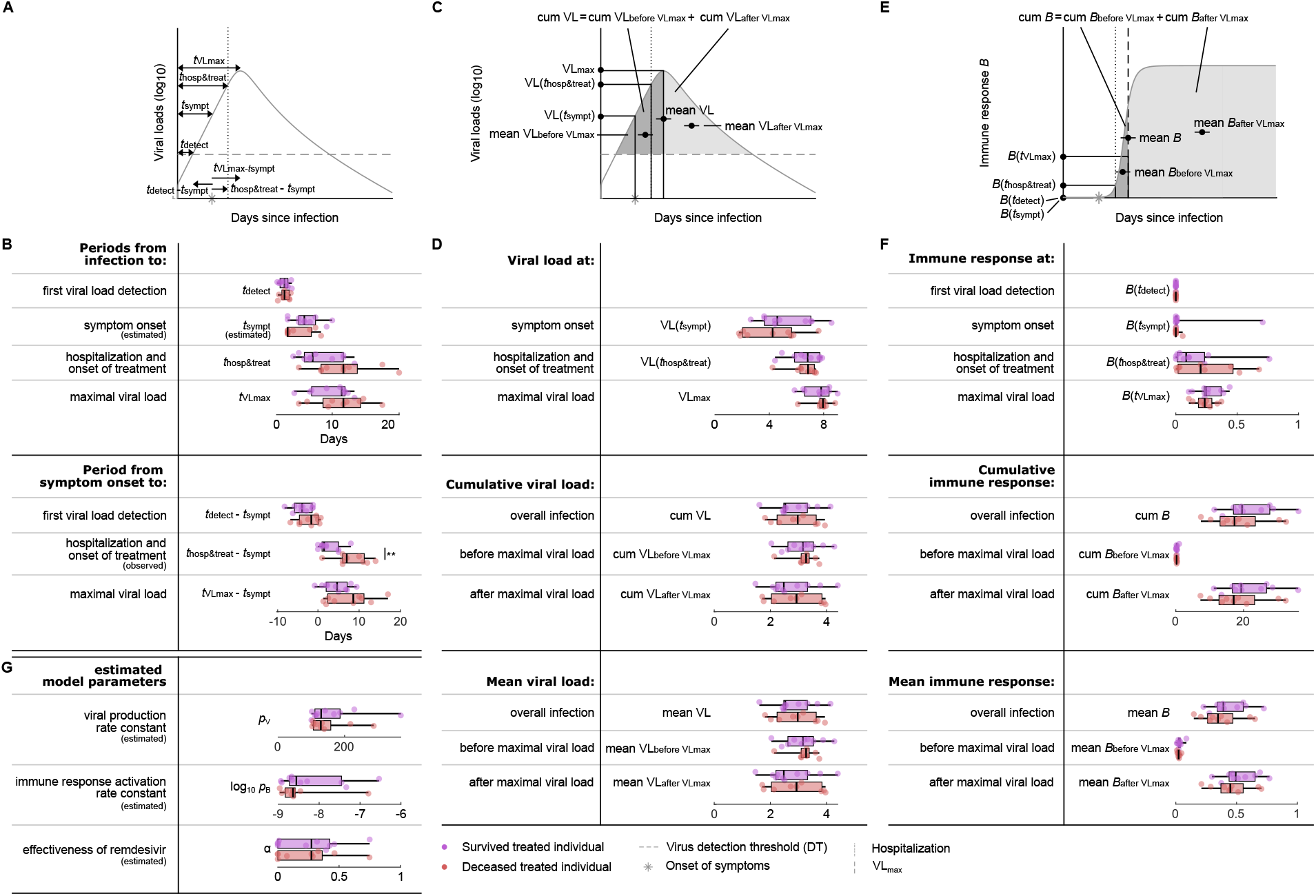
Clinical/virological timing, viral load, immune response features and model parameters for survived and deceased treated individuals. (A) Schematic presentation of timing features. (B) The observation, estimates and predictions of the timing features in A for survived (purple) and deceased (red) treated individuals. The boxplots represent the median, lower and upper quartiles, and minimum and maximum values. Periods are relative to time of symptom onset, so no boxplot is shown for the incubation period. All features are predictions, if not specified differently. (C) Schematic presentation of viral load features. (D) The individual predictions of the viral load features from C. (E) Schematic presentation of immune response features. (F) The individual predictions for the immune response features in E. (G) Estimated rate constants.

Most of the investigated features and estimated model parameters quantifying the viral load dynamics do not differ significantly between the survived and deceased treated individuals. However, our data suggests that deceased individuals were on average hospitalized and treated significantly later after symptom onset than survived individuals (7.0 days [95% CI: 1.0-14.0 days] vs. 1.5 days [95% CI: 0.0-8.0 days], respectively) (Figure 5B). As time of hospitalization and start of treatment coincide in this cohort, it is challenging to assess their relative importance. Despite the observed delay in hospitalization and start of treatment after symptom onset in the deceased treated individuals, the predicted cumulative and mean viral loads were not statistically different, indicating that viral load dynamics might not be a reliable predictor of mortality risk in this cohort (Figures 5D). The predicted immune response features appear to be consistently smaller for deceased treated individuals, suggesting a weaker overall immune response, however none of the differences were statistically significant. Overall, we estimated a low effectiveness of the antiviral treatment with remdesivir and found that effectiveness does not differ significantly between survived and deceased treated individuals (0.3 [95% CI: 0.0-0.7%] and 0.3 [95% CI: 0.0-0.7%] respectively, Figure 5G). Moreover, we did not find significant correlations between initiation of treatment after symptom onset and the effectiveness of antiviral treatment with remdesivir in either the survived or the deceased individuals (Pearson’s correlation coefficients -0.26 (*p*-value 0.47) and 0.35 (*p*-value 0.35), respectively, using the Student’s t-test).

### Antiviral treatment with remdesivir does not substantially affect viral load dynamics

Due to the non-random treatment allocation in our study cohort, comparing treated to untreated individuals to investigate the effects of antiviral treatment with remdesivir on viral load dynamics, might be prone to bias or confounding. We therefore leveraged the estimated individual-level effectivenesses of antiviral treatment remdesivir to stratify individuals into responders (those with an estimated treatment effectiveness above 0.1) and non-responders (those with treatment effectiveness below 0.1). For this, we assumed that non-responding individuals exhibit viral load dynamics equal to untreated hospitalized individuals with comparable disease severity. We found the proportions of responders to be similar between the groups of survived (4/10) and deceased (4/9) individuals with similar median effectivenesses in both groups (0.4 and 0.3, respectively). We next compared the observed, estimated, and predicted timing (Figure 6A and 6B), viral load (Figure 6C and 6D), and immune response (Figure 6E and 6F) features, and estimated model parameters (Figure 6G) between responders and non-responders (Extended Data Table 3).

**Figure 6.**
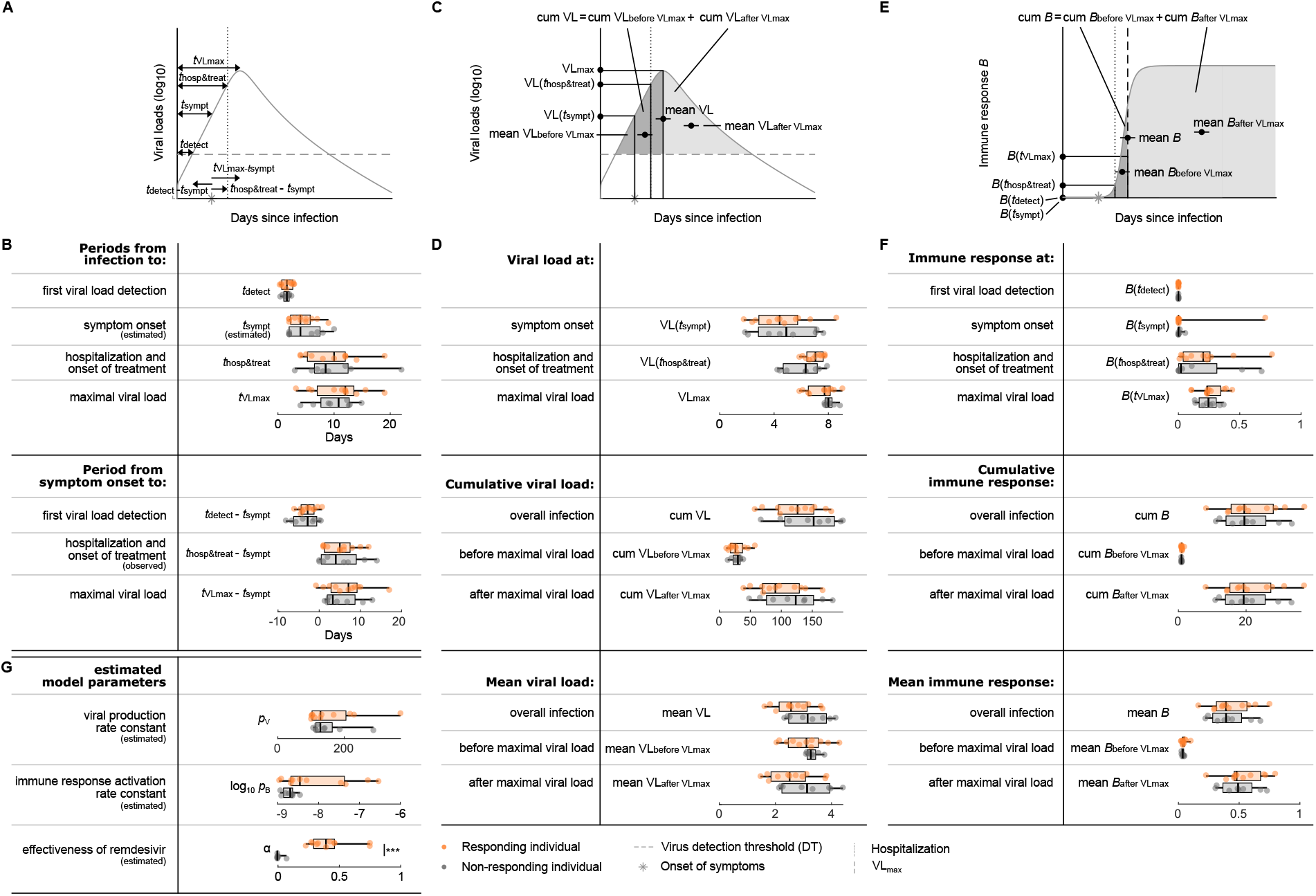
Clinical/virological timing, viral load, immune response features and model parameters for responding and non-responding individuals. (A) Schematic presentation of timing features. (B) The estimates and predictions of the timing features in A for responding (orange) and non-responding (gray) individuals. The boxplots represent the median, lower and upper quartiles, and minimum and maximum values. Periods are relative to time of symptom onset, so no boxplot is shown for the incubation period. All features are predictions, if not specified differently. (C) Schematic presentation of viral load features. (D) The individual predictions of the viral load features from C. (E) Schematic presentation of immune response features. (F) The individual predictions for the immune response features in E. (G) Estimated rates constants.

The effectiveness of antiviral treatment with remdesivir differed significantly between responders and non-responders with median values of 0.5 and 0.0, respectively (Figure 6G), demonstrating an effective discriminative data split. All other features were not significantly different between the two groups, suggesting that antiviral treatment with remdesivir fails to substantially reduce viral load, influence viral load dynamics, or shorten the time to undetectable viral loads. Moreover, there is no statistical evidence that treatment did notably mitigate mortality between responders and non-responders, with mortalities of 50% and 45%, respectively (*p*-value = 0.84, using the Chi-squared test). Collectively, our findings do not support a statistically significant effect of antiviral treatment with remdesivir on viral load dynamics.

## DISCUSSION

This study presents a comprehensive collection of virological, immunological, and clinical characteristics of a cohort of 369 individuals infected with SARS-CoV-2. In combination with mathematical modeling, our detailed analysis of 88 individuals offers important insights into the effects of hospitalization and antiviral treatment on viral load dynamics in both untreated and remdesivir-treated individuals.

The estimated and predicted timing and viral load features, such as the period from infection to viral load detection, incubation period, or the viral load at symptom onset, did not differ significantly between the hospitalized individuals with severe COVID-19 and the milder non-hospitalized ones (Figure 3). This result can be partially accounted for by the time discrepancy between most of the viral dynamics happening during early infection and the development of severe disease typically occurring later in the course of infection. It is further compatible with the well-known complex, multifactorial determination of clinical symptoms and disease severity in COVID-19, where within-host viral dynamics are only one factor alongside the immune response and other host-specific characteristics, such as age or the presence of a range of aggravating health conditions.

We found that the predicted cumulative viral load is significantly higher in the hospitalized than in the non-hospitalized untreated individuals, while the predicted mean viral loads are not significantly different between them (Figure 3D). This parameter combination indicates longer, more protracted infections in the hospitalized untreated individuals with otherwise similar viral load levels between individuals with severe and mild COVID-19. This finding aligns with previous reports [20] and reinforces the public health recommendation for prolonged isolation for individuals suffering moderate or severe COVID-19 [21]. Neither predicted cumulative viral loads, nor predicted mean viral loads were significantly different between treated individuals who died and those who survived, indicating that viral load dynamics are not a reliable predictor of mortality risk in this cohort (Figures 5D). Our model further indicates lower predicted immune response during early infection for hospitalized untreated individuals in comparison to the non-hospitalized untreated ones (Figure 3F). This result is compatible with both the identified longer duration of infection and the more severe disease in the group of the hospitalized untreated individuals [22].

We investigated the role of antiviral treatment with remdesivir on viral dynamics using model selection and statistical approaches. We first tested if treatment as a model parameter considerably improved the model’s ability to explain the viral load data of treated individuals. Model selection did not favor the model with treatment after accounting for its additional model complexity, suggesting that antiviral treatment with remdesivir does not considerably alter viral load dynamics in our cohort. To investigate the effect of remdesivir treatment on specific key virological and clinical outcomes, while controlling for potential biases stemming from the non-random treatment allocation in our cohort, we classified treated individuals into responders and non-responders as a proxy for treated and untreated individuals (see Results for details). In this, we assumed that non-responding individuals exhibit viral load dynamics comparable to those of untreated individuals with similar disease severity. The proportions of deceased responders and deceased non-responders are similar, suggesting that treatment does not influence mortality. Furthermore, our analysis did not find statistically significant differences in any of the investigated timing, viral load, and immune response features between the groups of responders and non-responders (Figure 6), suggesting that antiviral treatment with remdesivir fails to affect viral load dynamics. Finally, the median effectiveness of remdesivir between survived and deceased individuals did not differ, between these two groups, also suggesting a lack of impact of remdesivir therapy administration on patient survival. Collectively, our findings do not support any statistically significant effect of antiviral treatment with remdesivir on viral load dynamics or clinical outcomes.

Our results show that the median hospitalization and onset of treatment following symptom onset was considerably delayed in individuals who eventually died, compared to those surviving the infection (Figure 5B). This difference is substantial in magnitude and statistically significant. Survived treated individuals were hospitalized in the median 1.5 days after the onset of their symptoms, while it took a whole week for the deceased treated individuals to be hospitalized. As treatment was initiated at the same day as hospitalization, it is challenging to disentangle the relative importance of either using this data set. Yet, the low overall effectiveness of the antiviral treatment with remdesivir and the lack of difference between its effectiveness in survived and deceased individuals strongly points to the timing of hospitalization as the driving factor (Figure 5G). In contrast to previous studies highlighting the importance of early antiviral administration [18–20], we did not find significant correlations between the period from symptom onset to hospitalization/start of treatment and the effectiveness of remdesivir, underscoring the potential influence of other factors in determining treatment outcomes. Taken together, these findings suggest that early hospitalization post-symptom onset may bear greater significance for survival than antiviral treatment with remdesivir or than the timing of its initiation.

Our study suffers several limitations. We were unable to validate the model predictions for the immune response dynamics due to the lack of data. The size and scope of our sample did not permit us to stratify the data by typical confounding factors, such as age, sex, comorbidities, or viral load at admission while maintaining sufficient power for statistical inference [19]. We took steps to minimize potential variation in sample quality collection, targeting three independent SARS-CoV-2 sub-genomic regions for RT-qPCR (N/ORF1ab/S), and then accounted for measurement noise at the modeling stage. Finally, the large proportion of vaccinated individuals among the untreated non-hospitalized individuals may have further affected our overall results (Supplementary information).

In conclusion, our study provides important insights into SARS-CoV-2 infection dynamics in the context of antiviral treatment. Our results suggest a limited overall effectiveness of antiviral treatment with remdesivir on viral load dynamics and key clinical outcomes, while demonstrating the importance of early hospitalization for survival from COVID-19. Future work might focus on using similar procedures on evaluating the clinical effectivenesses of other antiviral drugs, such as nirmatrelvir or molnupiravir, and in particular combination drug therapeutic approaches, such as paxlovid [17,18]. With minimal need for adaptation, our modeling approach is amenable to future applications, with the important potential to provide insights into COVID-19 treatment strategies and the development of resistance that would not be possible to obtain using standard epidemiological approaches [17–19].

## METHODS

### Ethical statement

The research protocol was approved by the Research Ethics Committee of the Faculty of Medicine, University of Thessaly, Greece (84 / 09.12.2022), and was also approved by the Scientific Council of the public hospital from which the sample was derived (19535 / 28.06.2023).

### Study design

The 369 participants derived from individuals who tested positive for SARS-CoV-2 between October 2020 and September 2022. These individuals sought medical assistance at a public hospital emergency department situated in Larissa, Greece, due to exhibition of symptoms consistent with COVID-19 or close contact with a confirmed case of COVID-19. All of the participants had tested positive for SARS-CoV-2 based on nasopharyngeal/oropharyngeal samples. The gender (female/male) and age of each individual was recorded (Extended Data Figure 1). Moreover, we determined the specific variant of infection (alpha, beta, delta, or omicron), the status of COVID-19 vaccination (yes/no), previous COVID-19 infection(s) (yes/no), emerging symptoms (yes/no), comorbidity (yes/no), hospitalization (yes/ no), treatment (yes/no), admission to the intensive care unit (ICU) (yes/no), intubation (yes/no), and death from COVID-19 (yes/no). The cycle threshold (CT) was determined on at least 2 and up to 10 different days during the course of each infection using reverse transcriptase polymerase chain reaction (RT-PCR). The CT value denotes the number of PCR cycles required for the amplified viral load in the sample to cross the detection threshold (DT at CT = 37) and is hence inversely proportional to the viral load in the sample. We used the CT value as a proxy for viral load. Individuals were treated with antiviral treatment remdesivir according to the therapeutic protocols for COVID-19 of the National Public Health Organization of Greece. Antiviral treatment of hospitalized individuals was recommended for individuals with the need of oxygen therapy and mild COVID-19 or with the need of high-flow oxygen therapy and/ or non-invasive mechanical ventilation and/or severe COVID-19. Antiviral treatment was not recommended for individuals with clinical presentation lasting for more than 7 days, mild symptomatology and no need of oxygen therapy, and for individuals in extracorporeal membrane oxygenation or mechanical ventilation except for individuals to whom the administration of antiviral treatment had already been started.

### Laboratory analysis

Respiratory samples (nasopharyngeal or oropharyngeal swabs) were collected in transfer tubes containing viral transport medium and were analyzed in the Laboratory of Hygiene and Epidemiology, Faculty of Medicine, University of Thessaly. SARS-CoV-2 RNA was isolated from the samples with KingFisher Flex System (ThermoFisher Scientific, Waltham, MA, USA) using the MagMAX™ Viral/ Pathogen Nucleic Acid Isolation Kit (Applied Biosystems™, Waltham, MA, USA) according to the manufacturer’s instructions. Detection of the virus’s genetic material was performed using RT-qPCR with primers targeting SARS-Cov-2 specific genetic regions: ORF1ab, N, and S, with the TaqPath™ COVID-19 CE-IVD RT-PCR Kit (Applied Biosystems™, Waltham, MA, USA) on a validated QuantStudio™ 5 Real-Time PCR System (ThermoFisher Scientific, Waltham, MA, USA). The threshold for positivity ≤37 CT value for SARS-CoV-2 infection was established.

### Model cohort and pre-processing of individual viral load data

We focused our analysis on the subset of 90 individuals for which CT value measurements were taken for at least 4 time points during the SARS-CoV-2 infection. We extracted their personal ID numbers (in the format VOCXXX), measurement dates and corresponding CT values, date of symptom onset, whether the individuals were treated with antivirals (yes/no), were hospitalized (yes/no), and if so, the date of hospitalization, and, finally, whether the individuals died from COVID-19. We manually standardized the reported dates to the common date format MM/DD/YYYY. We next converted the dates at which the CT values were measured and if applicable, hospitalization, to days relative to the recorded individual-specific day of symptom onset. Missing CT values per target gene (N/ORF/S), denoted by --, were not accounted for. Negative results, denoted by NEG(-), were only reported if, and only if, all three target gene replicates were reported below the detection threshold of 37. For negative results we set the CT values of all three replicates to the detection threshold. We discarded one individual for whom the dates of CT value measurements were not recorded in a subsequent order and another individual who was treated but not hospitalized. In total, we considered 88 of the 369 individuals for the modeling analysis (Figure 1).

### Representativeness of subsets

To identify the features that are significantly over- or underrepresented in the subset of the model cohort with respect to the overall study cohort and the subsets of hospitalized and non-hospitalized untreated individuals and survived and deceased treated individuals with respect to the model cohort, we performed a statistical analysis (Extended Data Figure 1A and Supplementary information). For each feature and each subset, we simulated 10.000 sample subsets of X sample individuals, where X is the sample size of the respective subset, using MATLAB function *binornd(*X,*p,10000,1) [20]. p* is the success probability given by the fraction of the feature within the larger cohort (Extended Data Figure 1B). From the resulting 10.000 simulated sample subsets each of size X for a given feature, we calculated a point estimate and the 95% confidence interval. If the recorded fraction of the given feature of the subset is contained within the confidence interval, the feature is well represented by the subset. If the fraction of the given feature of the subset is below/above the confidence interval, the feature is under/ overrepresented by the subset. As an example: in the overall study cohort 56% of the recorded individuals were women. To identify whether the 55% of women within the model cohort are representative of the overall study cohort, we simulated 10.000 sample subsets of 88 individuals with a success probability *p* = 0.56. The resulting confidence interval with confidence level 95% is [45%, 66%]. As the recorded 55% of women in the model cohort is contained within the confidence interval, we term the fraction of women in the model cohort as representative of the fraction of women in the overall study cohort.

### Within-host SARS-CoV-2 model without treatment

The model comprises four populations corresponding to susceptible nasal epithelial cells *S*, infected nasal epithelial cells *I*, free virus particles *V*, and the immune response *B* (normalized to values between 0 and 1) (Extended Data Figure 2) [15]. The full system of nonlinear ordinary differential equations (ODEs) is given by the following equations:

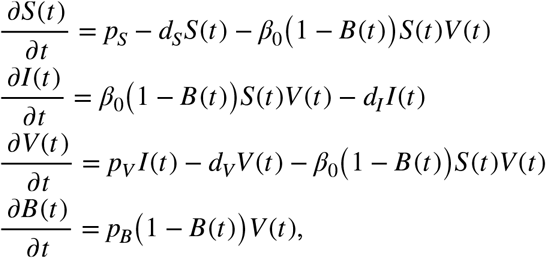

where *S*(0) = *S*_*0*_, *I*(0) = *I*_*0*_, *V*(0) = *V*_*0*_, and *B*(0) = 0 are the given initial values. Susceptible cells are constantly produced at rate *p*_*S*_ and die with natural death rate *d*_*S*_, where *p*_*S*_ = *d*_*S*_*S*_*0*_. When in contact with a free virus, susceptible cells get infected according to an overall infectivity rate *β*_*0*_(1-*B*(t)), where *β*_*0*_ is the infectivity rate. The term (1-*B*(t)) indicates the proportion of remaining successful infections not prevented by the immune response. The successfully infected cells enter the infected cell population and are eliminated with rate *d*_*I*_. Infected cells constantly produce and release free virus at rate *p*_*V*_, which is cleared at rate *d*_*V*_. Free virus leading to successfully infected cells are lost from this population according to the overall infectivity rate *β*_*0*_(1-*B*(t)). This type of approach allows for a global description of the immune response-virus interaction in tractable mathematical terms by maintaining a biological interpretation. We assumed the individual to not have been exposed to SARS-CoV-2 prior to this infection, hence, *B*(0) = 0. Upon infection, *B* increases with increasing viral load at rate *p*_*B*_ and is limited to a maximal relative strength of 1, such that *B*(t) ∈ [0, 1]. We considered the activation of the immune response to lead to a reduced overall infectivity rate *β*_*0*_(1-B(t)), and hence, to fewer successful infections of susceptible cells [13,21]. According to how the immune response *B* impacts the overall infectivity rate *β*_*0*_(1-*B*) in the model, an upper bound of *B* is necessary to avoid negative overall infectivity rates and, hence, to ensure biological meaningfulness. As this model is used to describe the short-term acute infection dynamics only, we ignore a decrease in the immune response to long-term post-acute infection levels. For more details on the full model accounting also for post-acute infection dynamics see [15]. Overall, the model is described by four populations, *S, I, V*, and *B*, and seven parameters, *p*_*S*_, *d*_*S*_, *β*_*0*_, *d*_*I*_, *p*_*V*_, *d*_*V*_, and *p*_*B*_, where all parameters are assumed to be positive for biological interpretability.

### Within-host SARS-CoV-2 model with antiviral treatment remdesivir

Remdesivir inhibits viral replication [22]. In this model, we assumed the viral production rate *p*_*V*_ to be compromised by a factor of 1-α, where α is the proportion of reduced viral production due to treatment and is termed effectiveness. Here, α ∈ [0, 1] (Extended Data Figure 2). If α = 0, then remdesivir treatment is completely ineffective, while α = 1 signifies a complete inhibition of viral production. We adjusted the third equation of the previous ODE system accordingly to

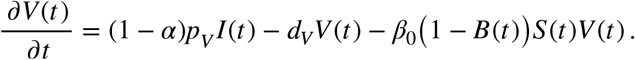

Remdesivir treatment was started at the day of hospitalization and was administered for five consecutive days. We assumed remdesivir treatment to be effective immediately and to be fully cleared after the last administration. After concluding the treatment, viral production is assumed to resume at its original rate *p*_*V*_.

### Parameterization of the within-host SARS-CoV-2 model without and with treatment

To describe the individual SARS-CoV-2 infection dynamics, we parameterized the model as shown in Extended Data Table 1. Initial values and the rate constants for 5 model parameters were taken from literature and assumed to be the same across individuals ([15] for more details). To account for the heterogeneity between individuals, we estimated the viral production rate *p*_*V*_ and the activation rate of the immune response *p*_*B*_ at an individual-specific level. Additionally, we estimated the incubation period, i.e., the duration from infection to reported symptom onset, *t*_*sympt*_ per individual. The upper and lower boundaries of *p*_*V*_ were determined by the range of estimated individual-specific parameters from Ke et al. [13]. The lower and upper boundaries of *p*_*B*_ are less interpretable and, hence, were assumed to cover a broad range of values, while the boundaries for *t*_*sympt*_ were set from 2 up to 14 days [23]. For the model with treatment, we also estimated the individual-specific effectiveness of remdesivir, which according to its definition is bounded between 0 and 1.

### Parameter estimation

To describe the transformed CT values using the model, we formally redefined population *V* as the virus in the sample and, similarly, *p*_*V*_ as the viral production rate times the proportion of sampled virus [13]. Accordingly, we also redefined *d*_V_, *p*_B_, and *β*_0_ times a factor reflecting the sampling process. To directly compare CT values and simulated viral loads, we made use of the CT value-to-viral load calibration determined by Ke et al. [13,23], given by

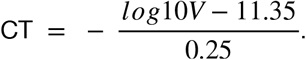

Experimental data such as the measurements of CT values is noise corrupt. We took this measurement noise into account in the model, by assuming an additive Gaussian measurement noise distribution. The log-likelihood for the Gaussian noise model for individual *i* time point *k* and replicate *j* with measured CT values 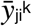 is given by

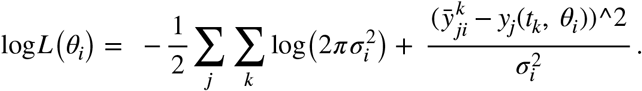

To account for the individual-specific measurement noise, we inferred an additional parameter σ determining the spread of the Gaussian noise model. The lower and upper bounds of σ were set to [10^-2^, 10]. In total, we estimated 4 parameters (*θ*_*i*_ = *p*_*V*_, *p*_*B*_, *t*_*sympt*_, σ}) for the untreated individuals (Figure 2) and 5 parameters (*θ*_*i*_ = *p*_*V*_, *p*_*B*_, *t*_*0*_, α, σ}) for the treated individuals (Figure 4). We then performed multi-start maximum likelihood optimization of the negative log-likelihood in the log_10_ parameter space for numerical reasons [24], initiating the optimization runs from 100 different Latin-hypercube-sampled starting points and maximizing over the CT values per individual. We used the MATLAB functions *fmincon* for optimization [25] and *ode45* for solving the ODEs [26].

### Model selection

For the 19 treated individuals, we fitted the model without and with treatment to the viral loads (Figure 4). To quantitatively compare the performance of both models and to determine the relative importance of treatment in the model, we used the Bayesian information criterion (BIC) [27]:

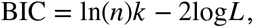

where *n* is the number of data points, *k* is the number of estimated parameters and log*L* is the log-likelihood value for the maximum likelihood estimate of the model parameters. The BIC rewards high likelihood values and penalizes additional model parameters. Hence, low BIC values are preferable. We considered a BIC difference of more than 10 between two models to be sufficient evidence to reject the model with the higher BIC [28] (Extended Data Table 4).

### Statistical comparison of timing, viral load, and immune response features and model parameters

We compared the distributions and medians of a total of 24, 29, and 29 timing, viral load, and immune response features and estimated model parameters between hospitalized and non-hospitalized untreated individuals, between survived and deceased treated individuals, and between treated individuals responding and not-responding to treatment. We used the two-sample Kolmogorov-Smirnov and Mood’s median tests as provided by the MATLAB functions *kstest2* [29] and *mediantest* [30] to compare the distributions and medians of the given features, respectively (Extended Data Table 3). As we were interested in every single feature separately, we did not correct for multiple testing.

### Implementation and code availability

The data and MATLAB code corresponding to this manuscript will be made available on GitHub upon acceptance of the manuscript. The analysis was performed with MATLAB 2023a.

## Data Availability

The data and MATLAB code corresponding to this manuscript will be made available on GitHub upon acceptance of the manuscript.

## ACKNOWLEDGMENTS

The views expressed are purely those of the authors and may not in any circumstances be regarded as stating an official position of the European Commission.

## AUTHOR CONTRIBUTIONS

Conceptualization, L.S., N.I.S.; Data curation, L.S.; Formal analysis, L.S.; Funding acquisition, N/A; Investigation, L.S., I.V., Z.B., V.A.M., C.H.; Methodology, L.S., N.I.S.; Project administration, N.I.S.; Resources, I.V., Z.B., V.A.M., C.H., N.I.S.; Software, L.S.; Supervision, N.I.S.; Validation, L.S.; Visualization, L.S.; Writing – original draft, L.S., N.I.S.; Writing – review and editing, L.S., P.V.M., N.I.S.

